# Consensus Guidelines for the Management of Peritoneal Surface Malignancies: Introduction and Methodology

**DOI:** 10.1101/2024.04.07.24305467

**Authors:** PSM Writing Group, PSM Consortium Group, Kiran K. Turaga

## Abstract

**Introduction:** The management of peritoneal surface malignancies (PSM) remains a challenge, characterized by limited high-level evidence and reliance on expert opinions. This paper outlines the methodology used in the development of clinical management pathways for PSM.

**Methods:** Building upon the Chicago Consensus Guidelines, a multidisciplinary North American panel was surveyed using a Modified Delphi technique to gauge the levels of agreement with clinical management pathways. Rapid systematic reviews were undertaken to address contentious questions specific to disease sites, which were structured using the Population, Intervention, Comparator, Outcome framework.

**Results:** Two consecutive rounds of voting were conducted between June 2023 and February 2024, addressing nine pathways spanning various conditions associated with PSM (colorectal, appendiceal, gastric, and neuroendocrine neoplasms, peritoneal mesothelioma, and gastrointestinal obstruction). The first Delphi consensus round involved 228 participants, of which 198 (87%) responded in the second round. Over 90% consensus was achieved in most blocks across all pathways. Eleven rapid systematic reviews were conducted, including 183 studies cumulatively out of 13,595 abstracts screened through PubMed. Perspectives from patient advocates and international experts were incorporated into these guidelines. The consortium’s multilevel approach also involved the creation of a peritoneal oncology curriculum, recommendations for billing and coding, and shared resources for perioperative care and patient information.

**Conclusion:** Experts indicated high levels of agreement with most pathway blocks across disease sites. These guidelines do not replace the need for high-level evidence but serve as guiding principles for critical decisions within the current landscape of PSM care.

## INTRODUCTION

Peritoneal surface malignancies (PSM) represent a significant challenge in oncology, characterized by a scarcity of high-level evidence, leading to management decisions often reliant on diverse provider opinions. To address these concerns, a multidisciplinary North American expert panel, herein referred to as the PSM consortium, convened to outline optimal care practices for patients with PSM. Expanding upon the previous Chicago Consensus Guidelines, the primary objective of the current process was to delineate consensus-based management pathways for various conditions associated with PSM.^1^ The evidence basis for these pathways were supported by rapid systematic literature reviews and insights from patient advocates and global leaders. This paper elucidates the methodology and limitations of the guideline development process.

## METHODS

### A. Pathways

#### Modified Delphi Consensus

To gather feedback regarding clinical management pathways, we employed a Modified Delphi method aligned with the Conducting and REporting DElphi Studies (CREDES) framework, with known advantages including leveraging collective expert opinions and the ability for virtual delivery.^2^ For this consensus, two rounds of voting were planned for all pathways after preliminary synthesis and review of major updates since the previous guideline iteration, with timelines depicted in Figure 1. Voting was performed on individual pathway blocks via electronic questionnaires, categorizing responses on a 5-point Likert scale ranging from “Strongly disagree” to “Strongly Agree” (Figure 2). The experts also provided reasons for disagreements on specific blocks and general comments. The consensus percentage was calculated based on the proportion of responses marked as “Strongly agree” and “Agree” among the total responses for a given block.^3^ A minimum consensus level of 75% was required to retain recommendations after the first round and blocks with < 90% consensus underwent further examination within disease site working groups After the first round of voting, we summarized the results and shared them with participating experts, allowing them to consider the group’s opinions before voting again. Only votes from members with complete responses in the first round were counted further in the second voting round, ensuring a proper subset. Responses from participants who voted in the second but not the first round were examined for comments, though not included in the reported Delphi round two consensus percentage.

**Figure 1.**
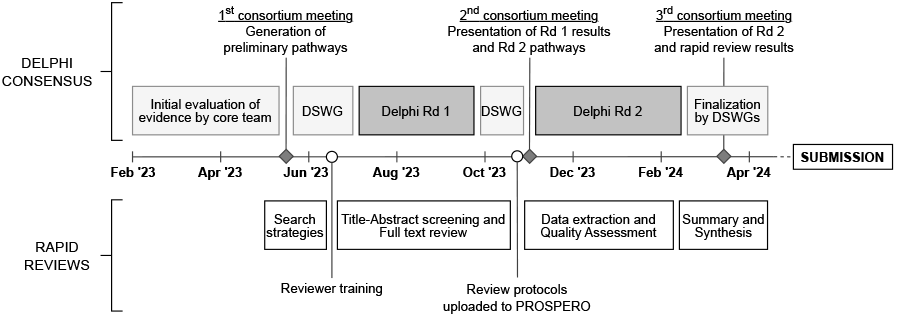
Timeline for the development of consensus guidelines for the management of peritoneal surface malignancies. Abbreviations: DSWG – Disease site working group, Rd – Round,

**Figure 2.**
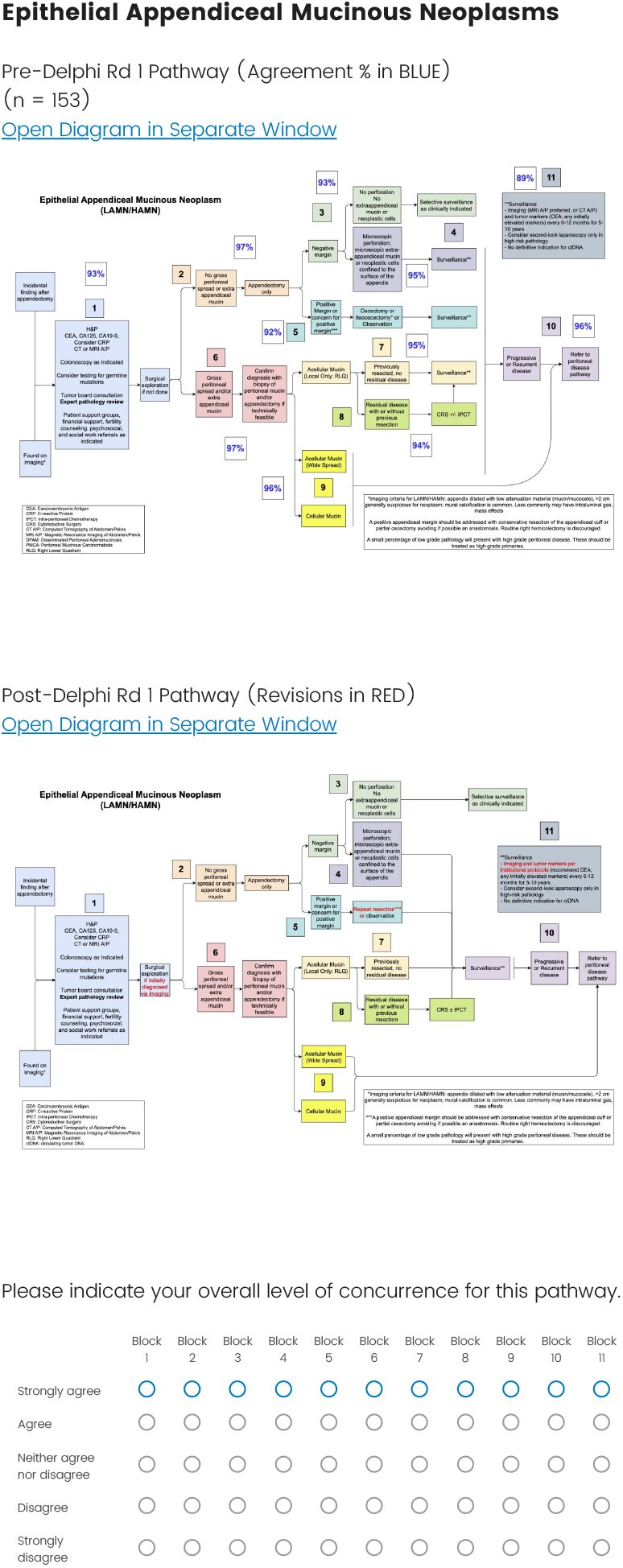
Survey example for the second Delphi consensus round presenting the consensus percentages for the first-round pathway version followed by the modified pathway, with changes identified in red text.

#### Selection of the Expert Panel and External Representatives

We engaged surgical and medical oncologists within the PSM care community and requested that they nominate colleagues from other disciplines. To expand our scope and engage people with varying levels of involvement in PSM care, we contacted members of the guideline panels from major national societies (NCCN, ASCO, and NANETs) via email.

Furthermore, we invited members of the Peritoneal Surface Oncology Group International (PSOGI) Executive Council to vote in the Delphi round two surveys and consolidated their responses separately to assess the alignment of our guidelines with global practices. Brief commentaries outlining salient similarities and differences compared to national and international guidelines were then synthesized for each guideline document. Additionally, we reached out to patient representatives through disease-site specific advocacy groups to incorporate their perspectives about clinical trial enrollment, research outcomes, and available resources in the context of PSM.^4,5^

#### Systemic Therapy and Pathology

During the first Delphi round, a need to streamline recommendations concerning systemic therapy associated with cytoreductive surgery and intraperitoneal chemotherapy across disease sites became evident. Consequently, we engaged medical oncologists within the consortium group to help create summary tables detailing first-line systemic therapies relevant to each disease site. Additionally, we engaged experts in gastrointestinal pathology within the consortium group to clarify the defining criteria for appendiceal tumors and pseudomyxoma peritonei in line with recent classification systems and their implications for treatment. Unlike the pathways, recommendations for systemic therapy and pathology were presented as summary tables alone and were not subject to formal voting.

### B) Rapid systematic reviews

To support the evidence basis for contentious matters arising during the pathway development process, we conducted a limited number of rapid systematic reviews in line with recommendations from the Cochrane Rapid Reviews Methods Group.^6^ PubMed was accessed through Medline as the primary search database. Key questions for these reviews were identified by disease site experts within the consortium and framed using the Population, Intervention, Comparator, Outcome, Design, Setting (PICO-DS) framework as detailed below.

- Population: Defining key clinicopathological characteristics of patients considered for inclusion, including the sample size.
- Intervention: Describing the nature and delivery of the interventions being studied.
- Comparator: Detailing attributes of the comparison group that often represent the standard of care.
- Outcomes: Specifying relevant health-related outcomes for studies of interventions and test parameters for studies regarding diagnostic tests.
- Design: Defining the study designs considered for inclusion. We excluded case series with a sample size of less than 10-20, case reports, and nonhuman studies.
- Setting: Considerations for the timing and choice of interventions, although sparingly used in our reviews as strict inclusion criteria.

The search strategies were subjected to peer review by a medical librarian specialist. Protocols for all reviews were developed a priori, verified by an epidemiologist, and registered in the PROSPERO online repository before data extraction (Figure 1). To streamline the review process, references were uploaded to Covidence, a screening and data extraction tool (Covidence, Melbourne, Australia). Abstracts and full-text manuscripts were reviewed in duplicate, and conflicts were resolved by a third reviewer. Excel spreadsheets were utilized for data extraction and quality assessment, which were performed as a single extraction with secondary verification. Quality assessment was performed using the Newcastle Ottawa Scale for nonrandomized studies and the Cochrane Risk of Bias (RoB) tool for randomized controlled trials.^7,8^ Summary tables were generated for each review and meta-analyses were conducted for questions that were feasible using random-effects models. Findings from the reviews are reported in disease site-specific documents aligning with the Preferred Reporting Items for Systematic Reviews and Meta-Analyses (PRISMA) guidelines.^9^

## RESULTS

Nine pathways were established: two for colorectal cancer, three for appendiceal tumors (mucinous neoplasms, localized appendix cancers, and peritoneal dissemination), and one each for gastric cancer, peritoneal mesothelioma, neuroendocrine neoplasms, and malignant gastrointestinal obstruction. Pathways addressing peritoneal dissemination from appendiceal tumors and gastrointestinal obstruction were newly developed, whereas the rest were updated from previous versions. Two pathways from the 2018 guidelines, focusing on desmoplastic small round cell tumors and ovarian cancer, were not further detailed in this consensus.

The first Delphi consensus round engaged 228 participants, with 198 (87%) responding in the second round. Tables 1 and 2 present the distribution of expertise and consensus percentages across disease sites, showing an increase in consensus percentages from the first to the second rounds across all pathways (Table 2). Eleven rapid reviews were conducted involving the screening of 13,595 abstracts and a review of 1,513 full texts, narrowing down to 183 articles included for data extraction and quality assessment (Figure 3).

**Table 1.** Delphi panel composition for the first round of voting.

| **Specialty** | **Colorectal** | **Appendix** | **Gastric** | **Mesothelioma** | **NENs** | **MGIO** |
| --- | --- | --- | --- | --- | --- | --- |
| Surgical oncology | 101 | 96 | 93 | 75 | 72 | 85 |
| Medical oncology | 25 | 20 | 16 | 13 | 18 | 14 |
| Pathology | 12 | 15 | 11 | 10 | 10 | 3 |
| Radiology | 1 | 1 | 1 | 1 | 3 | 1 |
| Palliative care | 1 | 2 | 1 | 1 | 0 | 4 |
| Patient/Caregiver group | 1 | 1 | 0 | 0 | 1 | 0 |
| Radiation oncology | 1 | 0 | 0 | 0 | 0 | 1 |
| Other | 3 | 3 | 2 | 1 | 3 | 3 |
| **Grand Total** | **145** | **138** | **124** | **101** | **107** | **111** |

Abbreviations: NENs – Neuroendocrine Neoplasms, MGIO – Malignant Gastrointestinal Obstruction

**Table 2.** Delphi Consensus results across all pathways (Attached)

|  | Colorectal<br>(Synchronous) |  | Colorectal<br>(Metachronous) |  | Appendiceal<br>Mucinous<br>Neoplasms |  | Appendix<br>Cancer |  | Appendix-Peritoneal<br>Disease |  | Gastric<br>Cancer<br>PM |  | Peritoneal<br>Mesothelioma |  | NENs |  | MGIO |  |
| --- | --- | --- | --- | --- | --- | --- | --- | --- | --- | --- | --- | --- | --- | --- | --- | --- | --- | --- |
|  | Rd 1 | Rd 2 | Rd 1 | Rd 2 | Rd 1 | Rd 2 | Rd 1 | Rd 2 | Rd 1 | Rd 2 | Rd 1 | Rd 2 | Rd 1 | Rd 2 | Rd 1 | Rd 2 | Rd 1 | Rd 2 |
| Block 1 | 98% | 99% | 97% | 99% | 93% | 99% | 96% | 99% | 96% | 98% | 98% | 98% | 95% | 98% | 96% | 99% | 97% | 98% |
| Block 2 | 95% | 99% | 96% | 98% | 97% | 98% | 94% | 97% | 83% | 94% | 97% | 96% | 94% | 98% | 92% | 98% | 81% | 96% |
| Block 3 | 94% | 98% | 94% | 99% | 93% | 96% | 91% | 98% | 96% | 98% | 97% | 97% | 95% | 97% | 89% | 95% | 94% | 98% |
| Block 4 | 77% | 88% | 88% | 99% | 94% | 98% | 96% | 98% | 88% | 95% | 96% | 99% | 91% | 99% | 93% | 98% | 97% | 97% |
| Block 5 | 94% | 99% | 92% | 97% | 92% | 98% | 97% | 99% | 93% | 97% | 89% | 88% | 88% | 96% | 94% | 98% | 86% | 94% |
| Block 6 | 92% | 96% | 94% | 98% | 97% | 99% | 96% | 99% | 96% | 98% | 83% | 85% | 94% | 97% | 93% | 97% | 93% | 98% |
| Block 7 | 93% | 99% | 92% | 97% | 96% | 98% | 90% | 95% | 93% | 95% | 91% | 96% |  |  | 86% | 99% |  |  |
| Block 8 | 82% | 93% | 90% | 96% | 95% | 98% |  |  | 82% | 91% | 92% | 99% |  |  |  |  |  |  |
| Block 9 | 93% | 96% | 86% | 99% | 96% | 96% |  |  | 88% | 98% |  |  |  |  |  |  |  |  |
| Block 10 | 86% | 96% |  |  | 96% | 99% |  |  | 77% | 95% |  |  |  |  |  |  |  |  |
| Block 11 | 88% | 99% |  |  | 91% | 95% |  |  |  |  |  |  |  |  |  |  |  |  |

Abbreviations: Rd 1- Round 1, Rd 2 – Round 2, NENs – Neuroendocrine Neoplasms, MGIO – Malignant Gastrointestinal Obstruction

**Figure 3.**
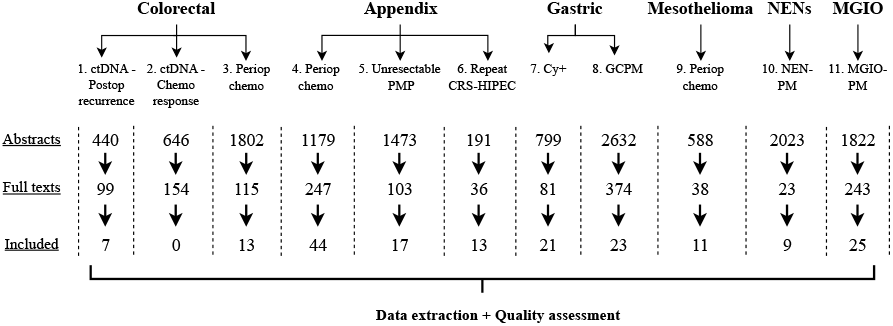
PRISMA flow diagram summary for all 11 rapid reviews. Abbreviations: ctDNA – Circulating tumor DNA, Chemo – Chemotherapy, Periop – Perioperative, PMP – Pseudomyxoma peritonei, CRS – Cytoreductive Surgery, HIPEC – Hyperthermic Intraperitoneal Chemotherapy, Cy+ - Cytology positive, GCPM – Gastric cancer peritoneal metastases, NENs – Neuroendocrine Neoplasms, MGIO – Malignant Gastrointestinal Obstruction

## DISCUSSION

These guidelines represent the collective efforts of over 250 individuals at outlining best care practices for managing PSMs. They substantiate the previous guideline iteration with a more stringent consensus and review methodology while engaging a larger spectrum of experts and patient advocates.^1010^ The evidence basis for decisions in PSM care primarily relies on heterogeneous retrospective data, as demonstrated in our reviews. It is crucial to acknowledge that not every clinical question can be addressed through a clinical trial; hence, leveraging consensus to streamline management pathways is pivotal to establish benchmarks for high-quality clinical care. This can pave the way for more uniform data collection methods, collaborative registries, and innovative clinical trials, which are long-term goals of our group.

Our approach extends beyond guideline development to encompass parallel efforts. The updated pathways were incorporated into an online mobile friendly PSM curriculum for medical students, residents, and fellows, designed in collaboration with program directors and pedagogical experts within the consortium group.^11^ Additionally, we surveyed surgical oncologists with expertise in PSM to assess institutional practices regarding billing and coding for CRS-IPCT procedures.^12^ Lastly, we established a repository of shared resources across institutions regarding perioperative care, Enhanced Recovery After Surgery protocols, and patient information materials, accessible through a common webpage.^13^

Our consensus and systematic review process had several limitations. There was an overrepresentation of surgical oncologists in the expert panel, which we attempted to mitigate by involving other disciplines more actively when reviewing feedback within disease site working groups and generating summaries for systemic therapies and pathology. Furthermore, the Delphi consensus involved voting on blocks rather than individual itemized recommendations. This approach was preferred to align with the original Chicago Consensus framework, although it may compromise the granularity of feedback received and overrepresent the consensus percentages for some disease sites. Owing to the anticipated heterogeneity of the data, only a limited number of rapid reviews were conducted per disease site to address contentious matters. Consequently, many recommendations in these guidelines are driven by expert opinion, which is considered to be low level evidence.^14^

Although these guidelines do not replace the need for higher levels of evidence, they aim to provide guidance for critical decisions within the current landscape of PSM care and a foundation for future collaborative efforts. With an additional focus on education, administration, and sharing resources, the PSM consortium aims to streamline the future of PSM management.

## Data Availability

The datasets used and/or analysed during the current study are available from the corresponding author on reasonable request. 

## ACKNOWLEDGEMENTS

We thank the Society of Surgical Oncology (SSO) and the Advanced Cancer Therapies Organization Committees for lending our group a dedicated meeting space during their annual conferences. We are grateful to the SSO Quality Committee for critically appraising the guidelines. We also thank the representatives from the PSOGI and patient advocacy groups for providing pertinent perspective commentaries. The included groups were as follows: Colontown, Appendix Cancer Pseudomyxoma Peritonei (ACPMP) Research Foundation, PMP Pals, Hope for Stomach Cancer, Mesothelioma Applied Research Foundation, and Learn Advocate Connect Neuroendocrine Tumor Society (LACNETS). We appreciate the inputs from Alexandria Brackett, a medical librarian specialist at the Yale Harvey Cushing Library, for examining the rapid review search strategies.

